# Systemic physiological “noise” in fMRI has clinical relevance

**DOI:** 10.1101/2025.08.19.25333215

**Authors:** Julia C. Welsh, Cole Korponay, Tianye Zhai, Justine A. Hill, Betty Jo Salmeron, Blaise B. Frederick, Amy C. Janes

**Author notes:** <u>Correspondence</u>: Amy Janes, National Institute on Drug Abuse (NIDA), Intramural Research Program, National Institutes of Health, Baltimore, MD 21224, USA.

## Abstract

Functional magnetic resonance imaging (fMRI) is central to studying neurobiological mechanisms, yet fMRI has limited clinical utility, highlighting the need for novel approaches. We show that a component of the fMRI signal—the systemic low-frequency oscillation (sLFO), linked to blood flow and physiological measures of arousal—indexes trait- and state-level drug use phenotypes. In individuals who chronically use nicotine, sLFO amplitude increased during abstinence and correlated with heightened dependence severity and cue-induced craving. In healthy participants, acute methylphenidate—but not nicotine—reduced sLFO amplitude in a manner that corresponded with improved behavioral performance. These findings demonstrate that the sLFO, typically treated as noise, carries biologically meaningful information. Evaluating the sLFO offers a complementary perspective to traditional fMRI analyses, thus enhancing clinical relevance. Broadly, the sensitivity of sLFO signals to drug administration, cues, and abstinence underscores the need to account for this signal’s contribution when interpreting fMRI responses across experimental conditions.

## Main

Functional magnetic resonance imaging (fMRI) is at the center of efforts to guide personalized, neurobiologically informed treatments for neuropsychiatric illnesses like substance use disorders. However, the current ability of fMRI data to explain or predict individual differences in drug use initiation, dependence severity, or treatment response remains underwhelming. The predominant use of fMRI in this context has been to gauge the interaction of drug use with neuronal activity and circuitry. However, drugs also interact with the brain’s vasculature. For instance, nicotine is a well-documented vasoconstrictor, and acute and chronic nicotine use affects vascular endothelial function and vascular tone and structure.^1^ Moreover, since the cerebrovasculature delivers drugs and their metabolites to and from neurons, intrinsic differences in cerebral hemodynamics may contribute alongside those in neurocircuitry to variance in phenotypes like dependence susceptibility. As such, the clinical utility of fMRI in the context of substance use disorders may be enhanced by considering the contribution of hemodynamics to variance in drug use phenotypes.

fMRI has the capacity to do this because the signal that it measures – the blood oxygen level-dependent (BOLD) signal – is a non-specific marker of cerebral hemodynamic activity. Typically, denoising strategies are used to isolate and study the neuronally-driven contributions to the fMRI BOLD signal. However, recent work has demonstrated that a component of the BOLD signal that is typically overlooked – the systemic low frequency oscillation (sLFO) – tightly reflects blood flow physiology. The sLFO, which occurs in the low-frequency range (∼0.009–0.15 Hz), is a pervasive global signal present throughout both the brain and body. In a direct comparison, the sLFO measured in brain-based BOLD data showed strong temporal overlap with the sLFO signal recorded simultaneously from a fingertip probe, confirming the sLFO’s widespread presence across both central and peripheral systems.^2^ Unlike low frequency oscillations in BOLD that are directly linked to local neuronal activity through neurovascular coupling,^3, 4^ which produce distinct, localized temporal patterns, the sLFO is a single global signal. The sLFO is found throughout the body, travels with the blood, and reaches different brain regions at different times due to varying delays in blood arrival time. This property, which reflects underlying vascular anatomy,^5^ allows the sLFO to be separated from the elements of the BOLD signal more linked with local neuronal activity.^6, 7^ The sLFO reflects fluctuations in blood oxygenation and volume that originate from physiological processes, such as heart rate variation, respiration, and vasomotion,^6^ which support effective tissue perfusion and clearance of interstitial fluid.^8^ Due to the link between the sLFO and physiology, the amplitude of the sLFO tightly mirrors physiological changes due to shifts in arousal.^9, 10^ While arousal itself is driven by neuronal processes, the sLFO does not directly measure this neuronal activity. Instead, the sLFO reflects arousal’s global effects on hemodynamics, and its impact on BOLD fMRI is further modulated by the time it takes for the blood carrying this signal to pass through the cerebral vasculature; this is in contrast to the direct *local* modulation of blood volume and oxygenation resulting from local neural activity as is typically assessed in traditional BOLD fMRI studies. While the sLFO may correlate with arousal, it is not generated by fluctuations in local neurovascular coupling, the basis of the traditional “neuronal BOLD signal.”^9–13^ Specifically, brain-wide average sLFO signal magnitude is inversely correlated with arousal level as measured by heart rate variability, respiration, and pupil diameter.^9, 10^ These findings align with previous work demonstrating that fMRI signal changes correspond with shifts in arousal measured both behaviorally and electrophysiologically,^11^ supporting the idea that certain components of the fMRI signal reflect arousal-related dynamics. However, such prior approaches have primarily relied on studies in non-human primates using template-based or EEG-linked methods.^11, 12^ Validating that the sLFO reliably correlates with clinical measures would offer a simpler, fMRI-only tool with strong potential for broad application in mental health research.

Despite its physiological relevance,^8^ the sLFO is typically viewed as noise.^9^ Critically, the sLFO can be removed without negatively impacting the evaluation of brain function. For instance, removal of the sLFO does not impact the ability to detect canonical functional networks,^2, 9, 13, 14^ proving that removal of the sLFO does not fundamentally disrupt well characterized brain signals. However, the dismissal of the sLFO as noise mirrors longstanding debates about the global signal in fMRI, which has similarly been treated as a noise despite evidence that it contains physiologically and behaviorally meaningful information.^15–18^ A growing body of research has demonstrated that fluctuations in the sLFO, global signal, and related physiological signals such as heart rate, respiration, and vasomotion, track changes in arousal, sleep state, anxiety, and even diurnal rhythms.^9, 10, 19–24^ While the global signal has been linked to arousal, its interpretation is complicated by the fact that it is comprised of not only the sLFO (summed over a range of delay times), but a mixture of noise sources, including head motion and scanner instabilities. In contrast, isolating the sLFO offers a cleaner and more targeted measure, making it a more reliable focus as a metric of physiology. Thus, rather than isolating the sLFO solely for the purpose of removing it as noise, this signal has the potential to be leveraged to study individual differences in physiological state that are behaviorally and clinically relevant— particularly in conditions where arousal and autonomic function are disrupted, such as substance use disorders.

While measuring physiology with other tools may be advantageous for non-neuroimaging research, in the context of fMRI studies, the sLFO can be extracted directly from the data, eliminating the need for additional physiological recordings, and allowing for retrospective analysis of prior data. Further, while the sLFO is related to systemic physiology, its utility lies not necessarily in interpreting it as a direct marker of arousal, but in recognizing its influence on BOLD signals and its potential to reveal meaningful physiological dynamics that correspond with clinically meaningful variables. While a wealth of research has focused on removing the sLFO to improve signal specificity,^13, 14, 25–31^ the current research moves the field forward by investigating the sLFO as a means to better understand the link between systemic physiological signals within the BOLD data and clinical features.

Here, we explore the potential of the sLFO to serve as a clinically relevant signal. Across four independent cohorts, we examine how the sLFO is impacted by 1) the presentation of salient drug cues to dependent users, 2) acute drug abstinence in chronic users, 3) acute drug administration in non-drug users, and 4) how such sLFO modulation relates to cognitive task performance. We also examine the extent to which the sLFO reflects more stable drug phenotypes like dependence severity. In the context of substance use, physiological arousal to stimuli such as drug-associated cues play a critical role in maintaining dependence,^32–36^ suggesting the potential clinical relevance of concurrently assessing brain-based measures of arousal and neuronal responding. While the current findings are specific to substance use, they translate to all fMRI research as they establish the general concept that the sLFO can be impacted by pharmacology and tasks, and that the sLFO relates to clinical measures and behavioral performance.

## Results

The clinical relevance of the sLFO was evaluated across four independent cohorts, using standard fMRI acquisition and pre-processing procedures. In all groups, the sLFO was modeled and quantified using Regressor Interpolation at Progressive Time Delays (RIPTiDe) analyses, which has been used to evaluate and remove sLFO signals.^6, 7, 9, 13, 14^

### sLFO Changes Across the Resting State and Cue Reactivity Scans

Our sample of nicotine dependent individuals (N = 64) were scanned 1.5 hours after smoking to standardize time since last cigarette use and fMRI data were captured at rest and during a smoking cue reactivity task. The cue reactivity task has been extensively used by our group and others^37–40^ and consists of five, 5-minute runs where participants are shown 10 smoking, 10 neutral, and 2 target images presented in a pseudorandom order where no more than two of the same picture occurs sequentially. This design allows us to measure the sLFO at rest, prior to any cue-related provocation and then to assess the sLFO over the course of repeated cue-presentations. The whole-brain averaged sLFO for rest and each of the five cue runs were calculated. There was a significant main effect of scan block on the brain-wide sLFO (η² = 0.47; F(5, 63) = 11.32, *p* = 0.0000001; Figure 1). Post hoc two-sided paired *t*-tests revealed that the brain-wide sLFO at cue block 1 was significantly lower relative to the sLFO at rest (t(63) = - 3.13; *p* = 0.002; d = −0.40; 95% CI [−0.66, −0.14]) and relative to any of the other cue blocks (Cue block 2: t(63) = −4.08, *p* = 0.0001, d = −0.37, 95 % CI [−0.56, −0.18]; Cue block 3: t(63) = - 5.32, *p* = 0.000001, d = −0.63, 95% CI [−0.88, −0.37]; Cue block 4: t(63) = −6.56, *p* = 0.00000001, d = −0.74, 95% CI [−0.1, −0.49]; Cue block 5: t(63) = −5.67, *p* = 0.0000004, d = −0.71, 95% CI [− 0.99, −0.43]). Cue block 2 was also significantly lower than cue block 4 (t(63) = −3.20; *p* = 0.002; d = −0.34; 95% CI [−0.55, −0.12]). No other significant differences were noted. These results indicate that the sLFO significantly drops during the first presentation of cues, which fits with the known inverse relationship between the sLFO and arousal—indicating that smoking cues initially increase arousal as indexed by a reduction in the sLFO. The sLFO then increases over the course of the cue reactivity task as the sLFO measured at cue block 1 was significantly lower than the other time points, suggesting a habituation of the sLFO over time. This finding indicates that the sLFO is dynamically influenced by environmental stimuli.

**Figure 1:**
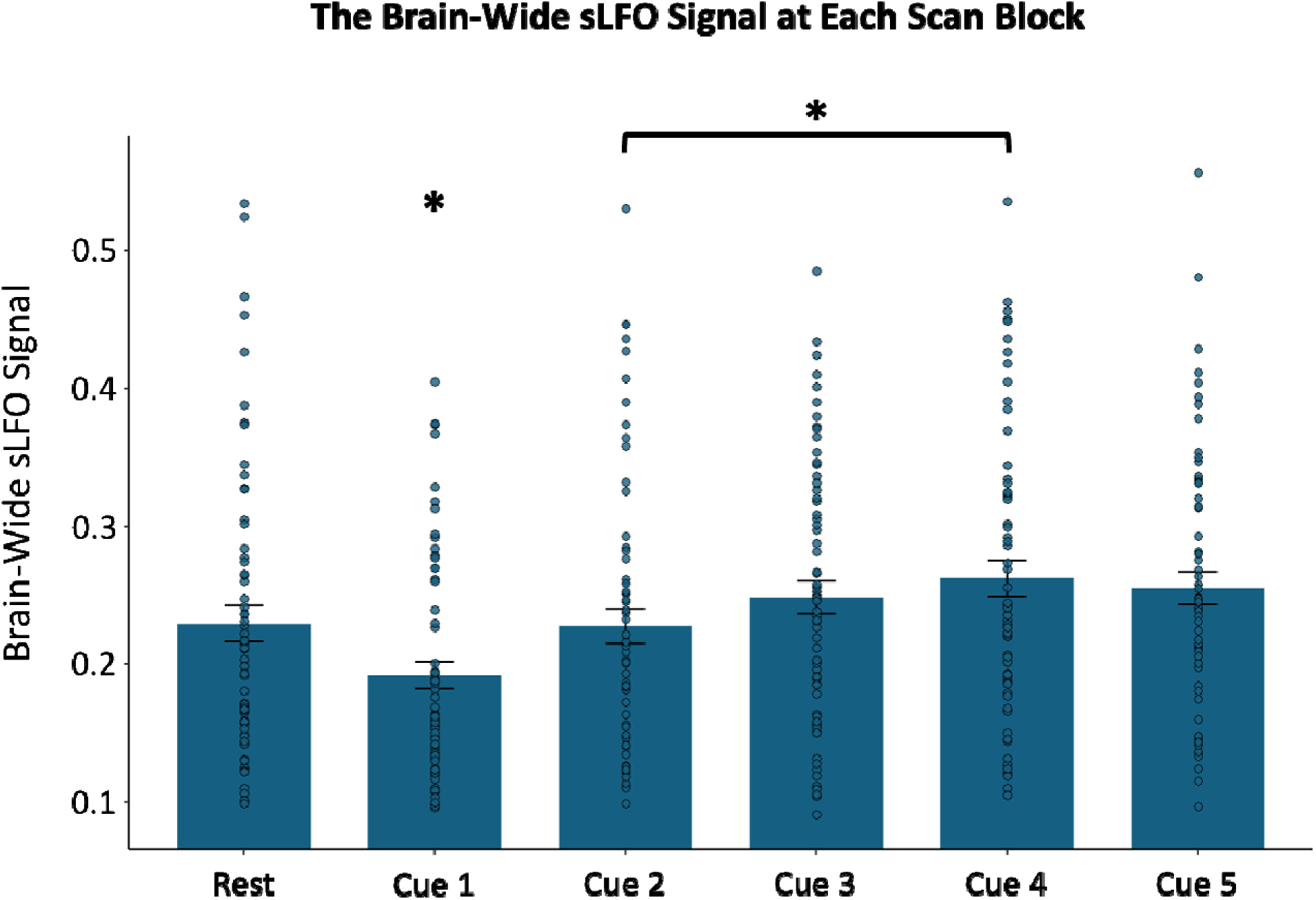
The brain-wide sLFO at rest and across the smoking cue reactivity task. The plot shows the significant main effect of scan block on sLFO (η² = 0.47; F (5, 63) = 11.32; *p* = 0.0000001). sLFO during cue block 1 was significantly lower than at rest (t(63) = −3.13; *p* = 0.002; d = −0.40; 95% CI [−0.66, −0.14]) and lower than during all subsequent cue blocks (Cue block 2: t(63) = −4.08, *p* = 0.0001, d = −0.37, 95 % CI [−0.56, −0.18]; Cue block 3: t(63) = −5.32, *p* = 0.000001, d = −0.63, 95% CI [−0.88, −0.37]; Cue block 4: t(63) = −6.56, *p* = 0.00000001, d = - 0.74, 95% CI [−0.1, −0.49]; Cue block 5: t(63) = −5.67, *p* = 0.0000004, d = −0.71, 95% CI [−0.99, - 0.43]). Cue block 2 also showed significantly lower sLFO compared to cue block 4 (t(63) = - 3.20; *p* = 0.002; d = −0.34; 95% CI [−0.55, −0.12]). No other pairwise comparisons between scan blocks were significant. Standard error bars are depicted for each scan block.

### The sLFO is Related to Nicotine Dependence and Craving

In the same cohort of 64 nicotine-dependent participants, we assessed relationships between the sLFO and clinically relevant measures of dependence severity and subjective craving. This builds on the prior finding which indicates that the sLFO can reflect state-related changes in arousal due to environmental stimuli. Here, we ask if such measures of the sLFO relate to smoking behavior and subjective experiences. Nicotine dependence was recorded as a static, trait-based measure using the Fagerström Test for Nicotine Dependence (FTND),^25^ where participants had an average FTND score of 4.77 ± 2.02, indicating moderate dependence. FTND scores were negatively correlated with the brain-wide sLFO averaged across all runs of the cue reactivity task (r = −0.324; 95% CI [−0.53, −0.09]; *p* = 0.009; Figure 2). At rest, the sLFO and FTND were unrelated (r = −0.047; 95 % CI [−0.37, 0.12]; *p* = 0.71). FTND and the sLFO at rest were also unrelated (r = −0.01; 95% CI [−0.21, 0.19]; *p* = 0.77) in an additional independent sample of 97 nicotine dependent individuals with an average FTND score of 4.59 ± 1.94 (moderate dependence). Collectively, these findings indicate that resting, or non-cue provoked measures of the sLFO, are unrelated to nicotine dependence severity. In contrast, it is those individuals with the lowest average sLFO—representing the greatest arousal—during cue presentation that have greater nicotine dependence severity. Not only is this in line with a wealth of literature linking the importance of drug cues in the maintenance of substance use,^33, 41–44^ but it also highlights the need to evaluate the sLFO under different contexts to fully appreciate its clinical relevance.

**Figure 2:**
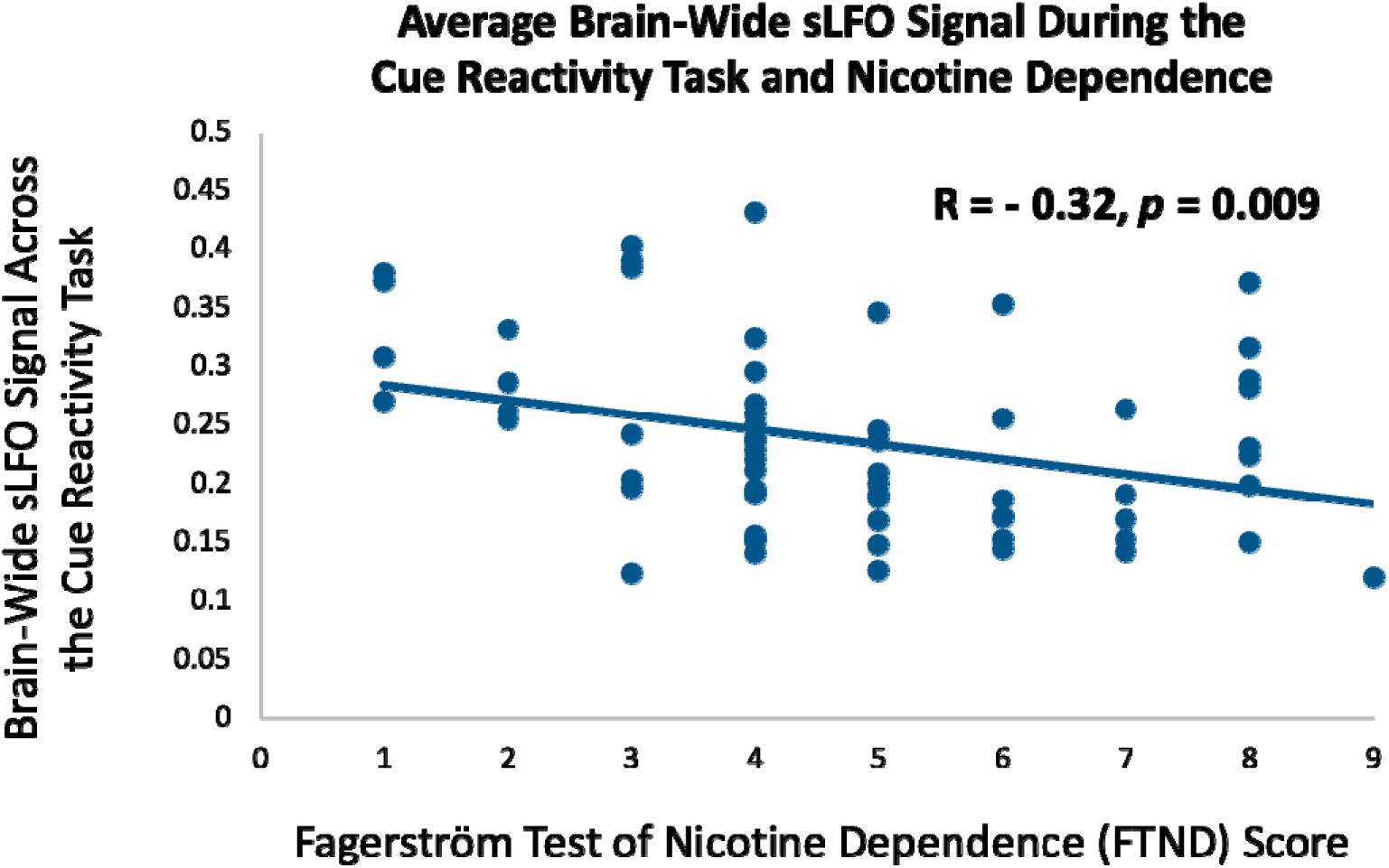
The relationship between nicotine dependence and the sLFO during the smoking cue reactivity task. FTND scores were significantly negatively correlated with the brain-wide sLFO across the five blocks of the cue reactivity task (r = −0.324; 95% CI [−0.53, −0.09]; *p* = 0.009), indicating that more severe nicotine dependence was associated with lower sLFO during the task. However, FTND scores were not significantly correlated with the sLFO at rest (r = - 0.047; 95 % CI [−0.37, 0.12]; *p* = 0.71).

We then asked whether the dynamic rise in craving typically seen following smoking cue exposure can be tied to changes in the sLFO across the cue reactivity task. Craving was measured using the Questionnaire of Smoking Urges (QSU) Brief Form,^45^ and cue-induced craving was calculated as: post-scan QSU – pre-scan QSU. Previously, we showed that such measures of subjective cue-induced craving increase following the cue reactivity task, and such changes in craving relate to neurobiology.^46^ Here, we found that greater increase in cue-induced craving was correlated with a smaller rise in the brain-wide sLFO across the cue reactivity task (r = −0.31; 95% CI [−0.51, −0.06]; *p* = 0.014; Figure 3). This indicates that those who experienced the greatest rise in craving following cue exposure maintained heightened arousal, as indexed by a smaller rise in the sLFO across the cue reactivity task. To confirm that this relationship was driven by the rise in sLFO over time and not by variance in the initial sLFO at cue block 1, the relationship between cue-induced craving and the cue-induced change in sLFO was calculated again, controlling for the initial sLFO during the first cue block. A similar relationship was found (r = −0.26; 95% CI [−0.48, 0.01]; *p* = 0.04), confirming the concept that changes in subjective cue-induced craving correspond with the brain-measured sLFO, which likely reflect variance in arousal responses to the cues.

**Figure 3:**
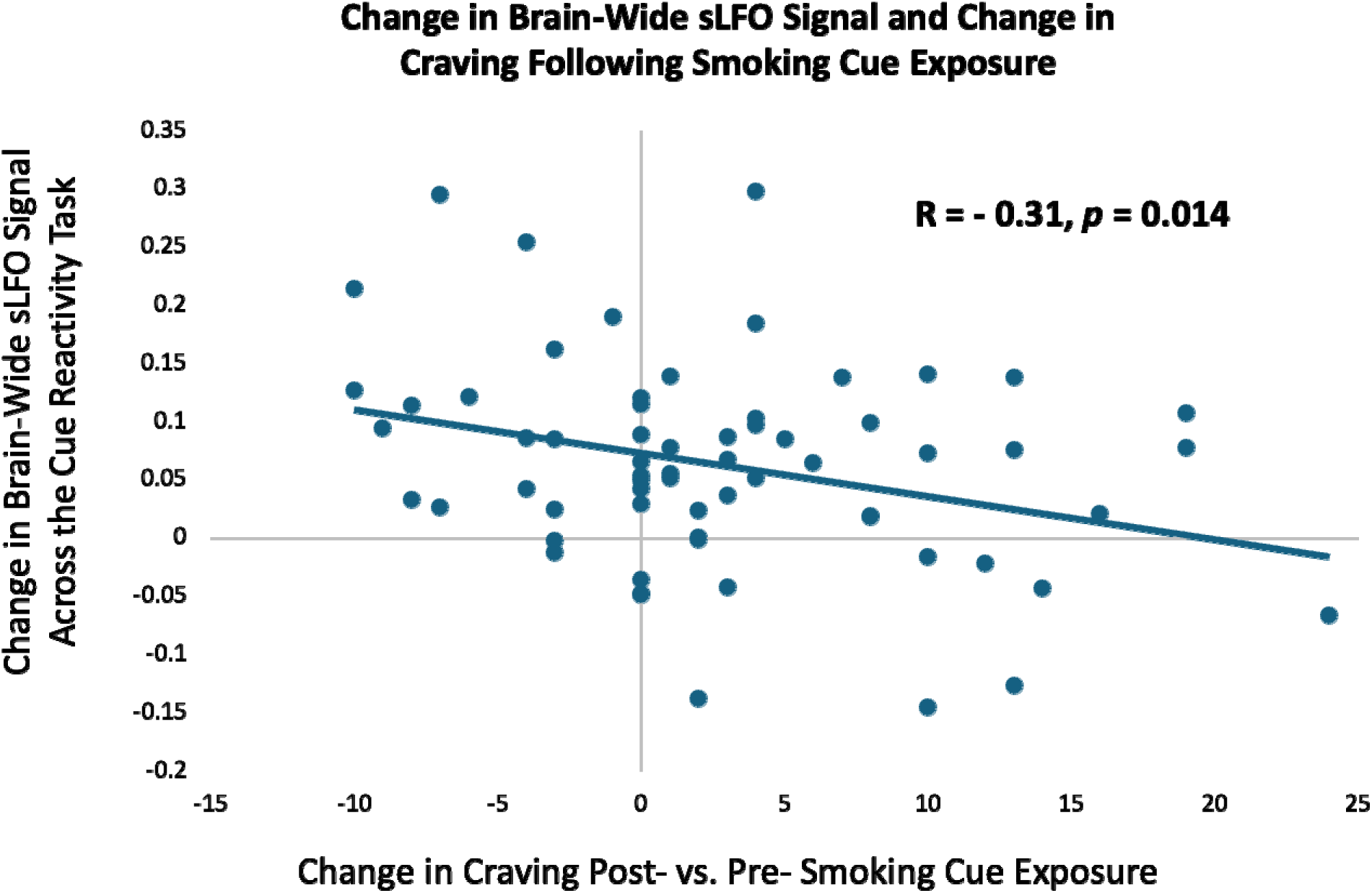
The relationship between cue-induced changes in subjective craving and the brain-wide sLFO. The magnitude of nicotine craving increase during the cue reactivity task (post > pre smoking cue exposure) was significantly negatively correlated with the change in brain-wide sLFO signal strength (cue block 5 sLFO – cue block 1 sLFO) (r = −0.31; 95% CI [− 0.51, −0.06]; *p* = 0.014).

### sLFO and Brain Reactivity to Smoking > Neutral Cues

The findings above center on the sLFO component of the BOLD signal instead of traditional measures of brain function. We have extensively used the cue reactivity task described here to assess cue-related brain function,^37–39, 47, 48^ yet it is unclear whether the sLFO has any impact on these more commonly used techniques. Therefore, we conducted a standard fMRI analysis looking at brain reactivity to smoking > neutral stimuli using BOLD data that included or removed the brain-wide sLFO from the BOLD data. Reactivity to smoking > neutral cues was significantly greater in brain regions including the medial prefrontal cortex (mPFC), posterior cingulate cortex (PCC), precuneus, and bilateral angular gyrus (Z > 3.1, *p*_corr_ = 0.05; Figure 4). This activation pattern is the same for data with and without the sLFO as the results overlap with r = 0.81. To further probe this finding and given prior work showing enhanced smoking > neutral reactivity in the anterior insula and default mode network (DMN)^49^, we conducted a follow-up region of interest (ROI) analysis evaluating the relationship between the brain-wide sLFO and cue reactivity in the insula and DMN using BOLD data with the sLFO removed (Figure S1).^47^

**Figure 4:**
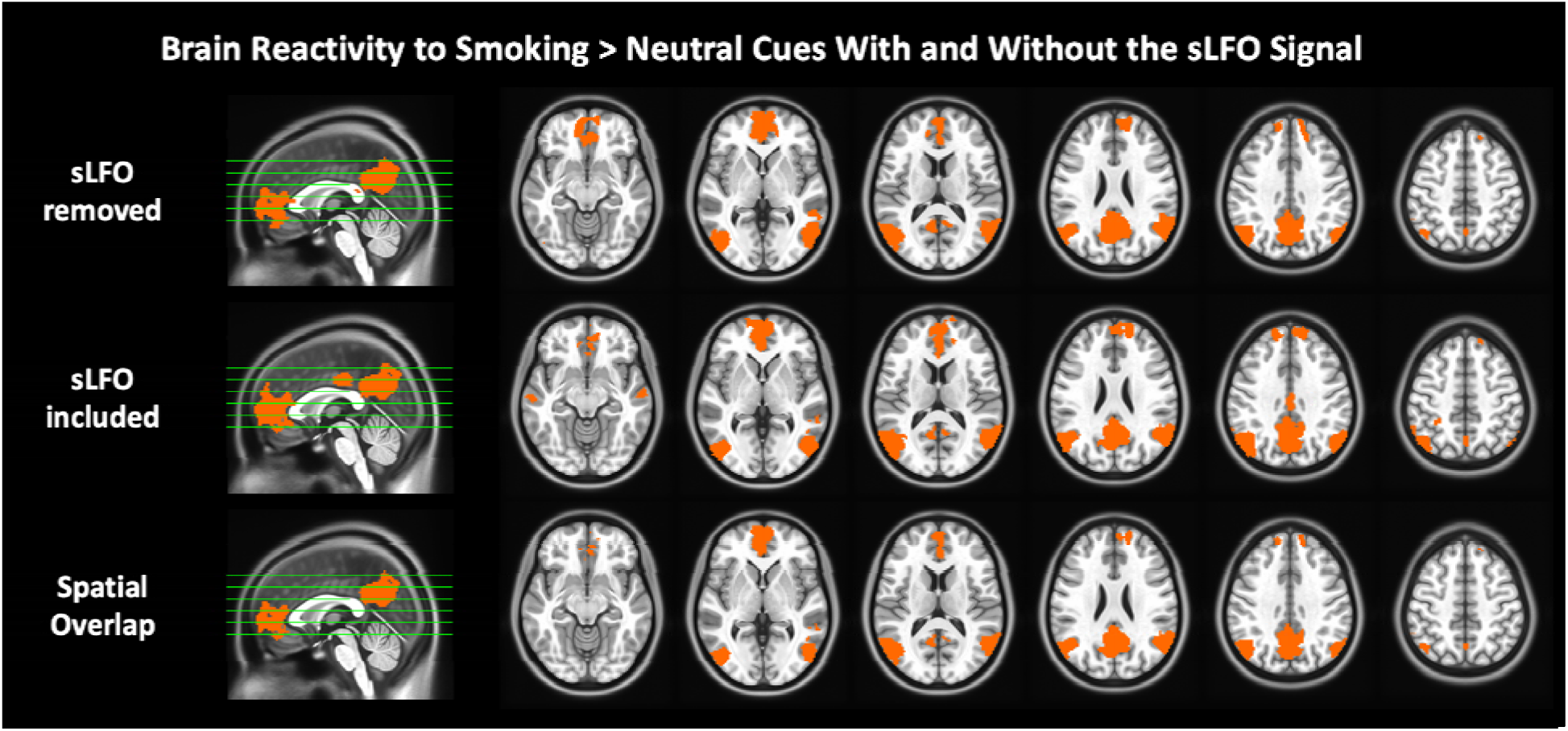
Smoking vs. neutral cue reactivity with the sLFO signal removed (top row), with the sLFO signal included (middle row), and their spatial overlap (bottom row). For both processing streams Z > 3.1, *p*_corr_ = 0.05, and activation maps overlap with r = 0.81.

There was no relationship between the brain-wide sLFO across the cue reactivity task and the smoking > neutral activation within the DMN (r = −0.18; 95% CI [−0.41, 0.07]; *p* = 0.259) or insula (r = −0.10; 95% CI [−0.34, 0.15]; *p* = 0.417). These findings indicate that the sLFO did not have a significant impact on the standard smoking > neutral contrast, nor did the sLFO relate to the level of smoking > neutral reactivity in brain regions typically activated by this task. Collectively, this suggests that the brain-wide sLFO and the neuronally based measures of smoking > neutral brain activation provide complementary information. Given our findings suggesting the sLFO habituates across the cue reactivity task, we tested for ROI changes in smoking > neutral activation over the five cue runs calculated using a repeated measures ANOVA. There was no change in the DMN (η² = 0.09; F (4, 63) = 1.55; *p* = 0.187) or insula (η² = 0.04; F (4,63) = 0.61; *p* = 0.66) reactivity to smoking > neutral cues over time, indicating that while the sLFO changes across the task, the more neuronally based brain reactivity to smoking cues remains relatively more stable.

### sLFO Increases During Abstinence Relative to Satiety

As the previously described findings link the brain-wide sLFO with cue reactivity, nicotine dependence, and craving, we asked whether those who do (N = 97) and do not use nicotine (N = 34) show differences in the sLFO. No significant difference was found between the sated nicotine-using group versus the healthy control group in the brain-wide sLFO (t(129) = −0.705; *p* = 0.482; d = −0.14; 95% CI [−0.54, 0.25]). To determine if there were more localized group differences in the sLFO, a voxel-wise comparison between these groups was also conducted and similarly no differences were noted. However, within those who chronically use nicotine, the brain-wide sLFO significantly decreased during nicotine satiety compared to abstinence (N = 65; t(64) = −4.51; *p* = 0.00003; d = −0.54; 95% CI [−0.79, −0.29]; Figure S2). Relative to abstinence, the voxel-wise sLFO significantly decreased during satiety in regions between the superior and inferior sagittal sinus, such as the medial primary motor cortex (mM1), and precuneus (*p_corr_* < 0.001, cluster size > 160mm^3^; Figure 5).

**Figure 5:**
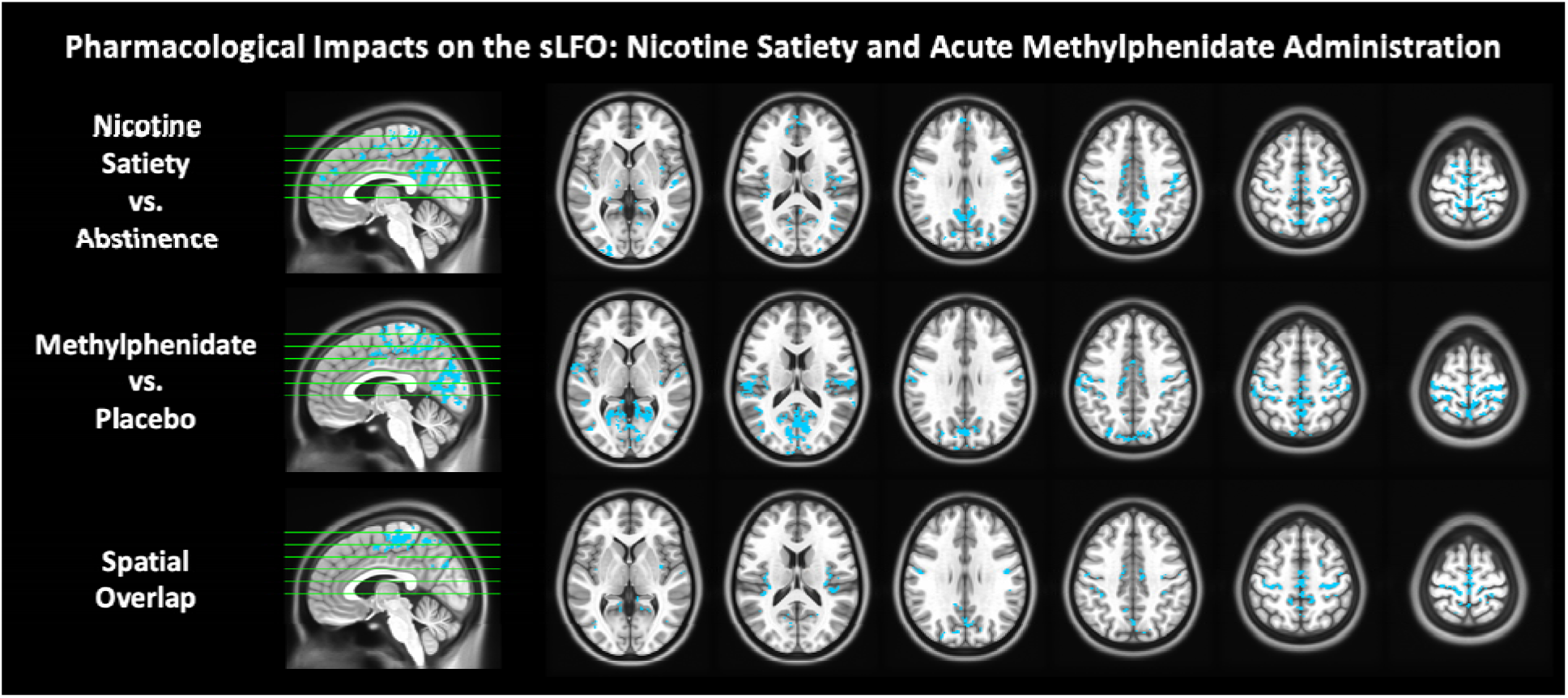
The voxel-wise sLFO signal at rest during nicotine satiety versus abstinence (top row), acute methylphenidate administration versus placebo (middle row), and their spatial overlap (bottom row). The voxel-wise sLFO signal significantly decreased during nicotine satiety relative to abstinence and during acute methylphenidate (MPH) administration relative to placebo. These reductions in the sLFO overlapped in regions between the superior sagittal sinus, inferior sagittal sinus, medial primary motor cortex (mM1), and precuneus (*p* < 0.001, cluster size > 160 mm³).

### Scan Order Does not Impact the sLFO

In the sample of nicotine dependent individuals scanned during satiety and abstinence, the abstinence visit was always conducted second to align with the larger clinical trial being conducted with this sample. To determine if there generally is an order effect on the sLFO, we used data from the Human Connectome Project (HCP) focusing on 462 young adults who we previously assessed as they have no current or family history of certain psychiatric illness or substance use (181 male, 281 female; age 28.66 ± 3.65 years). Critically, data in this group were captured on two independent days, with two scans per day recorded using different phase encode directions: right to left (RL) and left to right (LR). No significant difference in brain-wide sLFO was found between Day 1 and Day 2 when considering only the first scans (i.e., RL_1 and LR_2; t(455) = 0.11; *p* = 0.916; d = 0.01; 95% CI [−0.08, 0.09]) or second scans of each day independently (i.e., LR_1 and RL_2; t(455) = 0.62; *p* = 0.534; d = 0.03; 95% CI [−0.06, 0.12]). Upon collapsing the scans from Day 1 and Day 2 across RL and LR conditions, no significance difference emerged between the two days overall (t(1822) = 0.36; *p* = 0.716; d = 0.02; 95% CI [- 0.06, 0.10]). The voxel-wise sLFO confirmed this null finding, supporting the idea that the sLFO is not significantly impacted by scan order, which validates the idea that the sLFO is lower during satiety versus abstinence due to pharmacological changes.

### Effects of Acute Drug Administration on the sLFO

Since the sLFO was lower during satiety relative to abstinence in those who chronically use nicotine, we aimed to clarify nicotine’s pharmacological impact on the sLFO by administering a 7mg patch of nicotine to those who do not use nicotine (N = 58). Relative to placebo, the acute administration of nicotine had no impact on the sLFO at rest either at the brain-wide (t(57) = - 0.521; *p* = 0.604; d = −0.05; 95% CI [−0.29, 0.19]) or the voxel-wise level. It is plausible that this null finding is due to the fact that the 7mg dose of nicotine was too low to have an acute impact on the sLFO and that the finding in those who chronically use nicotine may be driven by physiological adaptations due to chronic use. However, it is plausible that other pharmacological manipulations directly influence the sLFO. To test this, the same participants received a 20mg dose of methylphenidate on a separate scan day, where methylphenidate is a psychostimulant known to increase dopamine and norepinephrine leading to increased arousal and vascular effects^50^. Compared to placebo, the administration of methylphenidate significantly decreased the brain-wide sLFO at rest (t(57) = −3.434; *p* = 0.001; d = −0.55; 95% CI [−0.88, −0.22]; Figure S3). This decrease was found to be driven by a reduction in the voxel-wise sLFO signal in brain regions between the superior and inferior sagittal sinus, such as the medial primary motor cortex (mM1), precuneus, and cuneus (*p_corr_* < 0.001, cluster size > 160mm^3^). These regional sLFO changes induced by methylphenidate overlap with the brain regions that demonstrate lower levels of sLFO during nicotine satiety relative to abstinence (Figure 5), indicating similarities between these two drug states on the sLFO.

Next, we tested whether there was a link between the sLFO and behavior; specifically determining whether pharmacological changes in the sLFO and pharmacological changes in task performance were related. The same cohort of non-substance using individuals who received separate acute doses of nicotine and methylphenidate as described above (N = 58) also performed the multisource interference task (MSIT) concurrently with fMRI. The brain-wide sLFO was extracted from the BOLD data captured during the MSIT task. The methylphenidate-induced decrease in the brain-wide average sLFO during the MSIT was associated with improved reaction times for both the congruent (r = 0.38; 95% CI [0.14, 0.58]; *p* = 0.003) and incongruent conditions (r = 0.39; 95% CI [0.14, 0.59]; *p* = 0.002) and increased accuracy for congruent (r = −0.60; 95% CI [−0.74, −0.40]; *p* = 0.0000009) and incongruent conditions (r = - 0.60; 95% CI [−0.74, −0.40]; *p* = 0.0000008; Figure 6). While there was no link between nicotine-induced changes in the brain-wide sLFO during the MSIT and reaction time for either the congruent (r = 0.07; 95% CI [−0.18, 0.34]; *p* = 0.578) or incongruent conditions (r = 0.08; 95% CI [−0.18, 0.34]; *p* = 0.527), there was an association between the nicotine-induced sLFO and improved accuracy for congruent (r = −0.35; 95% CI [−0.61, −0.18]; *p* = 0.006) and incongruent conditions (r = −0.39; 95% CI [−0.60, −0.17]; *p* = 0.002; Figure 6). These findings demonstrate links between pharmacologically-induced changes in both the sLFO and task performance, which is likely related to the association between the sLFO and arousal.

**Figure 6:**
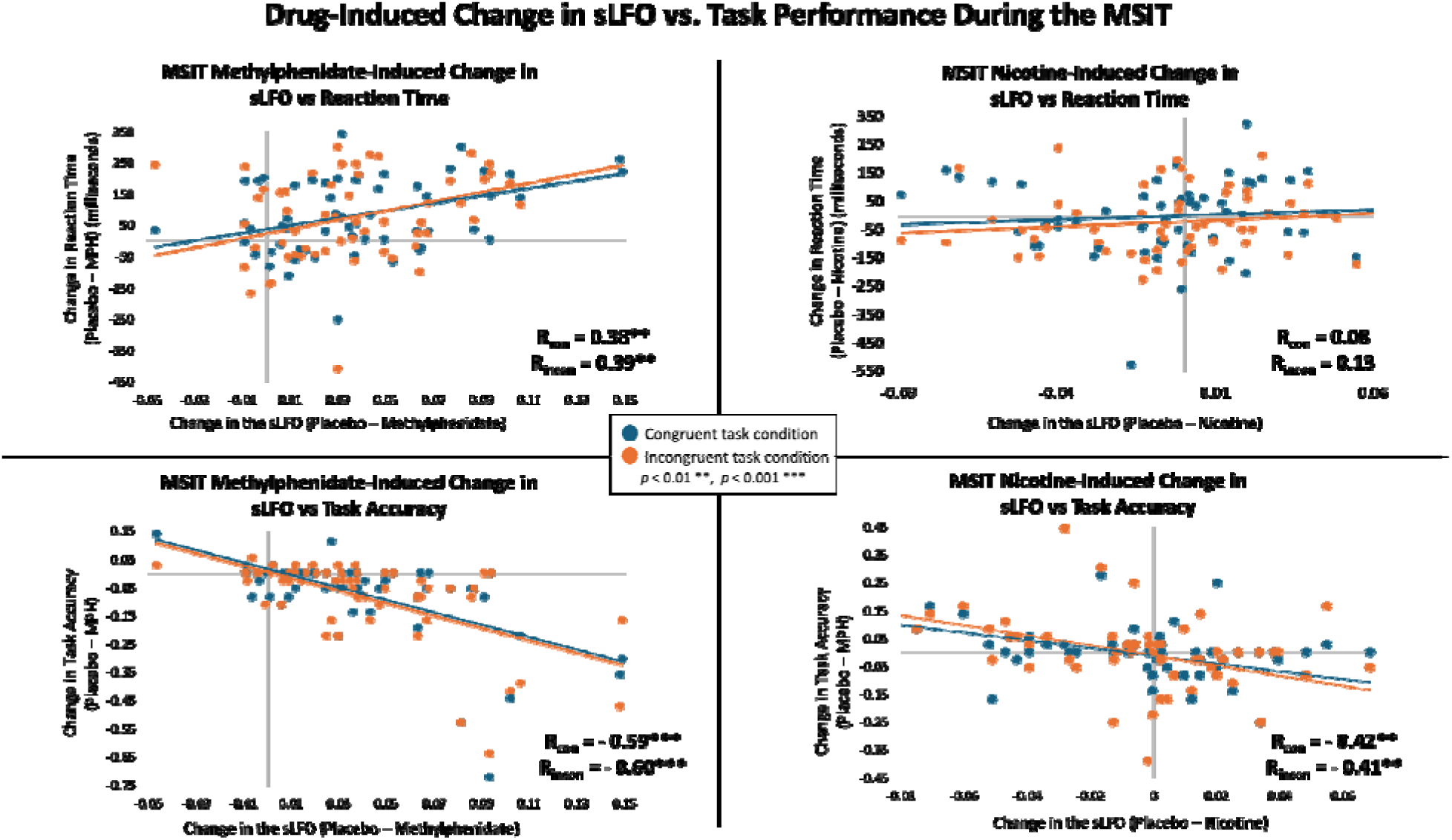
Correlations between the MSIT drug-induced change in the brain-wide sLFO versus reaction time and task accuracy in individuals who do not use nicotine. The methylphenidate-induced change in sLFO (placebo sLFO – methylphenidate sLFO) was positively associated with the difference in reaction time (placebo – methylphenidate) for both the congruent (r = 0.38; 95% CI [0.14, 0.58]; *p* = 0.003) and incongruent conditions (r = 0.39; 95% CI [0.14, 0.59]; *p* = 0.002) and negatively associated with the difference in accuracy (placebo – methylphenidate) for congruent (r = −0.60; 95% CI [−0.74, −0.40]; *p* = 0.0000009) and incongruent (r = −0.60; 95% CI [−0.74, −0.40]; *p* = 0.0000008) conditions. There was no relationship between nicotine-induced changes in sLFO and reaction time for either the congruent (r = 0.07; 95% CI [−0.18, 0.34]; *p* = 0.578) or incongruent conditions (r = 0.08; 95% CI [−0.18, 0.34]; *p* = 0.527). There was an association between nicotine-induced changes in sLFO and accuracy (placebo – nicotine) for congruent (r = −0.35; 95% CI [−0.61, −0.18]; *p* = 0.006) and incongruent conditions (r = −0.39; 95% CI [−0.60, −0.17]; *p* = 0.002).

## Discussion

This study shows that the sLFO component of the BOLD signal is clinically relevant. During the cue reactivity task, the brain-wide sLFO fell during the first cue block relative to the preceding resting scan. Our past work demonstrated that a *reduction* in the brain-wide sLFO amplitude represents *heightened* physiological arousal,^9^ which is in agreement with other work.^10^ Therefore, the sLFO reduction by the first set of cues fits with the known arousal-inducing effect of drug-related cues.^32–34, 51, 52^ Brain reactivity to smoking cues has been linked with aspects of substance use disorders including treatment outcome,^48, 53^ and subjective report of cue-induced craving is consistently linked with drug use.^40^ The current work indicates that the sLFO during cue reactivity also has clinical meaning as not only did cue exposure impact the sLFO, but the brain-wide sLFO during the cue reactivity task was negatively associated with nicotine dependence severity. Given that lower sLFO is linked with greater physiological arousal, this negative association suggests a relationship between more pronounced levels of physiological arousal during the task and heightened dependence severity. In contrast, there was no relationship between nicotine dependence and the brain-wide sLFO at rest, demonstrating that the link between nicotine dependence and the sLFO is context-dependent, only emerging when the sLFO is impacted by drug cue exposure.

Following the initial drop in the brain-wide sLFO, the signal rose over the course of the smoking cue reactivity task, indicating a habituation of arousal-related physiology over the task as a rise in sLFO is linked to *decreased* arousal. Critically, the rise in the sLFO across the cue reactivity task is negatively correlated with increased cue-induced craving (post – pre cue exposure). This negative association implies that, contrary to those who exhibit arousal-related habituation (greater rises in sLFO), those who maintain heightened arousal across the cue exposure task (smaller rises in sLFO) experience greater increases in cue-induced craving. Further, in contrast to the dynamic nature of the sLFO across the task, more neuronally based brain reactivity to smoking > neutral cues remained constant, indicating that brain reactivity to smoking cues persists despite sLFO habituation. Additionally, the whole-brain response to smoking > neutral cues was not changed when the sLFO was removed from the fMRI signal, and there was no relationship between the sLFO and insula or DMN reactivity to smoking cues. While not seen here, evidence suggests that under certain conditions, removal of the sLFO can improve activation map specificity^54^. However, such improvements are found in tasks with a single regressor where statistical significance is highly sensitive to both signal amplitude and baseline noise levels. In contrast, our cue-reactivity task models activation comparing conditions (smoking > neutral), which is more robust against inter-individual differences in baseline physiology (e.g., hematocrit, vascular responsiveness). Consistent with this, we observed only modest changes in activation patterns after sLFO removal, supporting the concept that contrast-based designs benefit less from sLFO removal as such task designs are less impacted by inter-individual physiology. This current modest change is not only expected due to our task design, but it demonstrates the reliability of task-related fMRI. Thus, fMRI data can provide two complementary sources of meaning to the study of mental health, one showing task-related activation and the second in providing physiological information as indexed by the sLFO.

Although there are links between the sLFO during cue reactivity and measures of craving and dependence, acute nicotine administration did not affect the sLFO at rest, and no differences were observed between individuals who do and do not chronically use nicotine. However, in those who chronically smoke nicotine, the sLFO was significantly reduced during satiety compared to abstinence. While the satiety scan was always conducted first, no scan-order effects were found in the large HCP cohort, indicating that the differences in the sLFO between satiety and abstinence reflect withdrawal-related pharmacological changes, rather than scan-order effects. Smoking and nicotine both have complex and extensive effects on vascular endothelial function, which alters vascular tone and structure, leading to cardiovascular disease.^1^ However, acute and chronic nicotine use may differentially impact the cerebrovascular system. For example, while acute nicotine use increases,^55^ chronic nicotine use impairs cholinergic angiogenesis.^1, 56, 57^ Therefore, it is plausible that acute and chronic nicotine use differentially impact the sLFO. While there is no direct evidence indicating that nicotine-induced changes in endothelial function significantly disrupts neurovascular coupling and thus the BOLD signal, such effects would not impact the current findings given the sLFO does not reflect local neurovascular coupling, but instead systemic vascular function. Additionally, the task design, which contrasts smoking > neutral stimuli is robust against inter-individual differences in physiology such as widespread alterations in neurovascular coupling. The lower regional sLFO observed during satiety, compared to abstinence, was found in brain regions between the superior and inferior sagittal sinuses, such as the medial primary motor cortex and precuneus. To test whether these effects generalize beyond nicotine, we included methylphenidate as a comparator drug. Both nicotine and methylphenidate are psychostimulants that modulate arousal and dopaminergic signaling, but through distinct mechanisms^50, 58, 59^—providing a broader view of the sLFO’s sensitivity to pharmacologically-induced changes in arousal. The voxel-wise sLFO was similarly impacted in overlapping brain regions following acute methylphenidate administration. These brain regions were predominantly located in sensory areas, overlapping with regions previously associated with physiological influences on the global signal.^29, 60–62^

During the MSIT, the methylphenidate-induced reduction in the sLFO was associated with faster reaction times and increased task accuracy, indicating that the drug-induced decreased sLFO was linked with a drug-induced improved task performance. While acute nicotine administration did not impact the sLFO at rest, the relationship between the nicotine-induced reduction in the sLFO during the MSIT and increased task accuracy mirrored that of methylphenidate. Collectively, these findings show a clear link between psychostimulant-induced reductions in the sLFO and improved cognitive performance, suggesting that studies evaluating the sLFO may aid in predicting behavioral consequences of pharmacological interventions.

The current work demonstrates the clinical relevance of the sLFO signal. Despite the strengths of this study, limitations should be noted. As discussed above, in our comparison of the sLFO during nicotine abstinence vs. satiety the scan order was fixed. This raises the possibility that the lower sLFO seen during the satiety scan could be due to scan order rather than pharmacological effects. To address this, we analyzed a large sample (N = 462) from the Human Connectome Project and found that scan order did not influence the sLFO. This suggests that our findings are unlikely to be due to scan order. Supporting this, we also observed lower sLFO in similar brain regions in both the nicotine (satiety vs. abstinence) and methylphenidate (methylphenidate vs. placebo) contrasts. This pattern indicates that psychostimulants may similarly influence the sLFO, further suggesting that the changes we observed during nicotine satiety were pharmacologically driven. Another limitation of the present study is that our work only examined nicotine and methylphenidate use. While more research is needed to generalize these findings beyond our sample and drug conditions, our results are likely applicable to other contexts. The current results show the sLFO is sensitive to both pharmacological interventions and emotionally evocative stimuli and the sLFO is linked to clinical variables and cognitive performance. Critically, these effects were consistent across different datasets, and they align with existing knowledge about the sLFO signal, supporting the idea that the findings will generalize to other populations.

In summary, the sLFO signal carries meaningful information. Unlike traditional physiological measures collected outside the scanner, the sLFO can be extracted directly from existing fMRI data without the need for additional equipment. This makes it a cost-effective, scalable tool for both retrospective and prospective studies. Given its sensitivity to pharmacological and emotional stimuli, future work should carefully consider the potential influence of such signals on interpretations of brain activity. While the sLFO is not itself a direct neural signal, its vascular characteristics may reflect the downstream physiological consequences of arousal-related neural processes. As such, it provides a complementary signal to those arising from local neurovascular coupling and contributes to the broader set of fMRI-based measures related to arousal and brain health. Isolating the sLFO not only improves the specificity of neuronal signals in fMRI,^9^ but also provides clinically relevant insights into pharmacological effects and other clinical variables. As such, integrating measures like the sLFO with traditional fMRI analyses may significantly expand the clinical utility of neuroimaging.

## Methods

Four cohorts were evaluated, with detailed descriptions included in the supplement. A brief overview is provided below. All participants were healthy and tested negative for pregnancy and acute drug/alcohol use (with the exception of nicotine where noted). All studies were approved by local institutional review boards.

### Participants

**Cue Reactivity Cohort** included 64 nicotine-dependent participants (39 male, 25 female; age 29.19 ± 6.93 years). Participants reported current nicotine dependence defined by the Fagerström Test for Nicotine Dependence (FTND; average 4.77 ± 2.02),^63^ and they had an average expired carbon monoxide (CO) concentration of 20.39 ± 12.51 ppm prior to scanning. To standardize the time between smoking and scanning, all participants smoked one of their own cigarettes 1.5 hours prior to scanning.

**Chronic Nicotine Use and Matched Controls Cohort** included 97 individuals who did (55 male, 42 female; age 48.23 ± 10.84 years) and 34 individuals who did not (22 male, 12 female; age 46.24 ± 9.45 years) chronically use nicotine. Participants in the nicotine-using group had an average FTND of 4.59 ± 1.94, and an average expired CO of 23.94 ± 11.96 ppm during their initial visit. Participants who did not use nicotine underwent one scan. Participants who used nicotine underwent at least one satiety scan while a subset of these individuals (N = 65) underwent a separate scan following 48 hours of nicotine abstinence (37 male, 28 female; age 47.85 ± 11.05 years). Abstinence was confirmed by a reduction in expired CO (2.63 ± 1.90 ppm; t(64) = 14.79, *p* = 2.5×10^−16^, d = 2.23, 95% CI [1.67, 2.79]).

**Human Connectome Project (HCP) Cohort** included 462 participants from the Young Adult dataset who we previously assessed given they have no current or family history of certain psychiatric illness or substance use^9, 64^ (181 male, 281 female; age 28.66 ± 3.65 years). Data were captured on two independent days, with two scans per day recorded using different phase encode directions: right to left (RL) and left to right (LR).

**Acute Drug Administration Cohort** included 58 healthy control individuals (17 male, 41 female; age 31.17 ± 9.37 years). Participants were required to have no lifetime history of substance use disorders and be free from any substance abuse for at least two years prior to participating. Participants were scanned following administration of 7 mg nicotine, 20 mg methylphenidate, or placebo – timed in accordance with drug peak – using a randomized, counterbalanced, double blind design.

### fMRI Acquisition

In all cohorts, fMRI data were collected at rest. fMRI data were also collected during a cue reactivity task within the Cue Reactivity Cohort only and during a Multi-Source Interference Task (MSIT) within the Acute Drug Administration Cohort only. Acquisition and preprocessing pipelines are provided in the supplement.

### Cue Reactivity Task

The cue reactivity task has been described previously.^37, 38^ Briefly, the task consisted of five, 5-minute runs where, for each run, participants were shown 10 smoking, 10 neutral, and 2 target images in a pseudorandom order, with no more than two of the same picture type occurring sequentially. Images were presented for 4 s followed by a 6-14 s (mean = 10 s) jittered intertrial interval consisting of a black screen and a white fixation cross. Smoking images included tobacco cigarette-related content such as people smoking, holding cigarettes, or cigarettes alone. Neutral images were matched in content and composition, involving people, hands, or objects but without smoking cues. Target images were animals and were included to ensure attentiveness via a button press. Subjective nicotine craving was measured before and after the task using the Questionnaire of Smoking Urges (QSU) Brief Form.^45^ To determine cue-induced craving, participants’ change in craving score was calculated as: post-scan QSU – pre-scan QSU.

### Multi-Source Interference Task

Participants were shown a three-digit number and were asked to indicate, via button-press in the scanner, the position of the number that differed from the other two items presented,^65^ this information was used to calculate accuracy and reaction time of responses. During congruent trials, the identity and position of the target numbers is consistent. During incongruent trials, the identity and position of the target numbers is inconsistent. The task consisted of 3 blocks, with each block consisted of 24 stimuli split evenly between trial types (12 congruent and 12 incongruent). Stimuli were presented for 2 s following by a 4-8 s jittered intertrial interval in increments of 500 ms.

### Data analysis

#### Systemic Low Frequency Oscillation (sLFO) Identification

In all cohorts, the sLFO was modeled and quantified using Regressor Interpolation at Progressive Time Delays (RIPTiDe) (https://github.com/bbfrederick/rapidtide). This method isolates low-frequency fluctuations (0.009–0.15 Hz) in the BOLD signal and estimates voxel-wise blood arrival times via cross-correlation to create lag maps that capture the temporal propagation of the sLFO across the brain.^6, 9, 13, 66^ These lag maps reflect vascular transit delays and are distinct from neural hemodynamic responses, thus proving that the sLFO is vascular in nature. For a more detailed discussion of lag maps, see Aso et al., 2017^65^ and Aslan, Hocke, & Frederick, 2025^67^.

A refined sLFO reference regressor is then generated by iteratively bootstrapping from the LFO-filtered global mean BOLD signal. This process uses cross-correlation to determine the arrival times of the sLFO, aligning voxel time courses, and applying principal component analysis to extract the reference regressor. We then created voxel-specific regressors by adjusting the refined sLFO regressor according to the blood arrival times for each voxel. This then provided, for each subject and scan condition, a map of the average sLFO amplitude for each voxel. In subsequent analysis, the reported sLFO findings refer to the average brain-wide sLFO amplitude across all brain voxels, whereas the voxel-wise sLFO specifically refers to the regional sLFO signal. A linear regression model was used to filter, fit, and eliminate the signal from the fMRI data, removing physiological noise without causing spurious negative correlations between regions, as previously shown.^13^ In the Cue Reactivity Cohort, fMRI data was used to evaluate the smoking > neutral contrast where the sLFO signal had been removed from the data and it was compared to data where the sLFO signal had been left in the data.

#### Cue Reactivity Task Analyses

The cue reactivity analysis was conducted using the FMRIB software library (FSL) (www.fmrib.ox.ac.uk/fsl). Following preprocessing, the first-level analysis was conducted on each of the participant’s five runs of the cue reactivity task. The first-level general linear model included regressors convolved with the gamma hemodynamic response function corresponding to smoking, neutral, and target images. Confound regressors modeled motion (*x*, *y*, *z* translation and rotation) and spikefix-identified artifacts. The smoking > neutral image contrast was conducted. First-level results were then combined (across the five task blocks) using a second-level fixed effects analysis to generate the average brain reactivity for each individual participant. The group-level analysis was then conducted using FLAME (FMRIB’s Local Analysis of Mixed Effects) to identify average brain reactivity across all participants and was corrected for multiple comparisons using a cluster threshold of Z = 3.1, *p* < 0.05. This analysis was independently conducted once after RIPTiDe analyses were applied to remove the sLFO signal and once before sLFO signal removal. The correlation between the smoking > neutral activation patterns for analyses with and without the sLFO signal were calculated using FSL’s FSLCC command.

Given prior work showing enhanced smoking > neutral reactivity in the anterior insula and default mode network (DMN),^49^ we conducted follow up region of interest (ROI) analyses evaluating the insula and DMN from data with the sLFO removed (Figure S1).^47^ ROI changes in smoking > neutral activation over the five task runs were calculated using a repeated measures ANOVA. A Pearson’s correlation was used to assess the relationship between the average ROI activation and average sLFO signal across the entire task.

#### Change in the sLFO Across Resting and Cue Reactivity Runs

A repeated measures ANOVA comparing the brain-wide sLFO was run considering scan block (rest, cue blocks 1-5). Post-hoc two-sided paired *t*-tests were conducted to define directionality of effect.

#### Relationship Between the sLFO, Nicotine Dependence, and Cue-Induced Craving

Separate Pearson correlations were performed between FTND and the sLFO amplitude measured: 1) during rest and 2) averaged across the five blocks of the cue reactivity task. To assess dynamic changes, we also tested the association between cue-induced craving (pre- vs. post-cue reactivity) and cue-induced change in the sLFO, calculated as the difference in the brain-wide sLFO between cue block 5 and cue block 1. Given that the FTND reflects a static measure of nicotine dependence, whereas cue-induced craving reflects a dynamic change in craving, we correlated FTND scores with the average sLFO across the five task blocks, and cue-induced craving with the cue-induced change in sLFO.

#### sLFO, Chronic Nicotine Use, Withdrawal, and Acute Nicotine and Methylphenidate

In the Chronic Nicotine Use and Matched Controls Cohort, the brain-wide sLFO at rest between conditions (individuals who used nicotine chronically vs individuals who did not use nicotine) was compared using a two-sample, two-tailed *t*-test. In the subgroup of individuals who used nicotine chronically, the brain-wide sLFO at rest during nicotine abstinence vs satiety was compared using a paired-samples two-tailed *t*-test. In the Acute Drug Administration Cohort, the brain-wide sLFO was analyzed separately at rest and during the Multi-Source Interference Task between conditions. The placebo vs acute nicotine/methylphenidate administration conditions were compared using paired-sample two-sided *t*-tests. To parse out regional differences in the voxel-wise sLFO at rest between conditions, paired-sample two-sided *t*-tests were conducted using whole-brain voxel-wise sLFO amplitude maps. Statistical significance was defined at voxel-level *p* <0.001 and randomization based multiple comparison correction at cluster level α <0.05, NN1 (face-wise nearest neighbor) (*3dttest++*, AFNI).

#### Potential Confound of Scan Day Order

In the Chronic Nicotine Use and Matched Controls Cohort, the sated and abstinent conditions were not randomly counterbalanced across scan sessions, yielding a potential confound of scan order. To test whether the sLFO changes as a function of scan day order, we conducted Day 1 versus Day 2 analyses of the brain-wide and voxel-wise sLFO strength in healthy control subjects in the HCP Cohort. Given that the HCP collected two scan sessions per day using different encoding directions (RL and LR), one-sided paired *t*-tests were conducted to assess within-subject differences between both scans on Day 1 versus Day 2. Differences in encoding direction were also evaluated. Second, a voxel-wise analysis was performed by calculating sLFO difference scores between the scans on Day 2 (averaging RL_2 and LR_2) and Day 1 (averaging RL_1 and LR_1). This difference was then tested using a 1-sample one-sided *t*-test using FSL randomize with 5000 permutations, applying a threshold of *p* < 0.05 with Threshold-Free Cluster Enhancement (TFCE).

#### Multi-Source Interference Task

The MSIT behavioral data analysis was conducted by calculating reaction times on correct trials and response accuracy during the congruent and incongruent conditions. The drug effect on the sLFO and task performance were calculated by subtracting the placebo values from drug condition values for the brain-wide sLFO, reaction time, and accuracy. The drug-induced change in brain-wide sLFO was then correlated with drug-induced task changes.

## Supporting information

Human Connectome Project (HCP) Cohort Accession Numbers

## Data Availability

Data collected at McLean Hospital and the National Institute on Drug Abuse (NIDA) can be made available per request and after obtaining a data transfer agreement. Data from the Human Connectome Project is publicly available on the open access Connectome database (https://db.humanconnectome.org/app/template/Login.vm). The specific HCP participant ID numbers are listed in the supplement.

## Acknowledgements

This work was supported by R01DA039135, the Intramural Research Program of the NIH/NIDA, and the NIDA Neuroimaging Research Branch.

This research was supported in part by the Intramural Research Program of the National Institutes of Health (NIH). The contributions of the NIH authors were made as part of their official duties as NIH federal employees, are in compliance with agency policy requirements, and are considered Works of the United States Government. However, the findings and conclusions presented in this paper are those of the authors and do not necessarily reflect the views of the NIH or the U.S. Department of Health and Human Services.

## Code Availability

The code and instructions for performing RIPTiDe denoising using the rapidtide package are publicly available at https://rapidtide.readthedocs.io/en/latest/usage_rapidtide.html#removing-low-frequency-physiological-noise-from-fmri-data.

