## Supplement for "Systemic physiological “noise” in fMRI has clinical relevance"

### Participants

**Cue Reactivity Cohort.** Clinical Trial Name & Number: Electronic Cigarettes and Reactivity to Smoking Cues, NCT01782599

The analyses presented here focus on data collected during the pre-treatment baseline assessment when all participants were still regularly smoking prior to the session. Participants were excluded if they met criteria defined by the Diagnostic and Statistical Manual of Mental Disorders (Fifth Edition) for moderate-severe drug or alcohol dependence, recent major depressive disorder (past 6 months), history of psychotic or bipolar disorder, serious medical illness, or pregnancy. All participants tested negative for current drug or alcohol use (excluding nicotine), and females tested negative for pregnancy. All procedures were completed at Harvard Medical School's McLean Hospital and were approved by the Partners Human Research Committee. Accrual began in October of 2015 and completed in April of 2021.

**Table S1.**

**Demographic Characteristics of the Cue Reactivity Cohort.**

| **Demographics** | |
|---|---|
| | **Individuals Who Use Nicotine (N = 64)** |
| Age (years) | 29.19 (*SD* = 6.93) |
| Sex (male/female) | 39/25 |
| Race (*n*) | |
| Black/African American | 6 |
| White/Caucasian | 47 |
| Asian | 7 |
| More Than One Race | 4 |
| Ethnicity (*n*) | |
| Hispanic | 6 |
| Non-Hispanic | 58 |
| Education (years) | 14.99 (*SD* = 2.01) |

Demographic characterization of the Cue Reactivity Cohort.

**Chronic Nicotine Use and Matched Controls Cohort.** Clinical Trial Name & Number: Imaging Biomarker for Addiction Treatment Outcomes, NCT03427424

All participants were recruited for a study investigating differences between individuals who did and did not use nicotine, with those in the nicotine-using group being offered enrollment in an individualized, counselor-involved treatment study following their initial study visit. No participants reported current psychiatric or neurological disorders as assessed by the Diagnostic and Statistical Manual of Mental Disorders (Fourth Edition). All participants tested negative for current drug or alcohol use (excluding nicotine for those in the nicotine-using group), and females tested negative for pregnancy. All procedures were completed at the National Institute on Drug Abuse and were approved by the institutional review board of the National Institutes of Health. Accrual began in October of 2013 and completed in June of 2023.

**Table S2.**

**Demographic Characteristics of the Chronic Nicotine Use and Matched Controls Cohort.**

| **Demographics** | |
|---|---|
| | **Individuals Who Use Nicotine (N = 97)** |
| Age (years) | 49.23 (*SD* = 10.84) |
| Sex (male/female) | 55/42 |
| Race (*n*) | |
| Black/African American | 47 |
| White/Caucasian | 43 |
| Asian | 4 |
| Native Hawaiian/Other Pacific Islander | 1 |
| More Than One Race | 2 |
| Ethnicity (*n*) | |
| Hispanic | 4 |
| Non-Hispanic | 93 |
| Unknown or not reported | 0 |
| Education (years) | 13.41 (*SD* = 2.64) |
| | **Healthy Controls (N = 34)** |
| Age (years) | 47.24 (*SD* = 9.45) |
| Sex (male/female) | 22/12 |
| Race (*n*) | |
| Black/African American | 21 |
| White/Caucasian | 12 |
| Asian | 0 |

| | |
|---|---|
| Native Hawaiian/Other Pacific Islander | 0 |
| More Than One Race | 1 |
| Ethnicity (*n*) | |
| Hispanic | 1 |
| Non-Hispanic | 32 |
| Unknown or not reported | 1 |
| Education (years) | 13.56 (*SD* = 2.00) |

Demographic characterization of the Chronic Nicotine Use and Matched Controls Cohort (N=131; N=97 individuals who use nicotine, N =34 healthy controls).

**HCP Cohort**.

All participants had no prior history of major psychiatric, neurological, or medical disorders known to affect brain function. Individuals were excluded from the analysis if they reported a family history of schizophrenia, met DSM-IV criteria for alcohol dependence, or had a lifetime history of significant substance use, defined as more than ten instances of cocaine, hallucinogen, opiate, sedative, or stimulant use; more than 20 instances of tobacco use; or more than 100 instances of marijuana use. Additionally, participants provided a breath sample indicating a blood alcohol content of less than 0.05 on the day of the scan, and a urine sample that tested negative for drugs of abuse (cocaine, marijuana, opiates, amphetamines, or methamphetamines). All procedures were approved by the Institutional Review Board at Washington University in St. Louis. Data collection took place at Washington University in St. Lous, Missouri. Accrual began in August of 2012 and completed in March of 2015.

**Table S3.**

**Demographic Characteristics of the HCP Cohort.**

| **Demographics** | |
|---|---|
| | **Healthy Controls (N = 462)** |
| Age (years) | 28.66 (*SD* = 3.65) |
| Sex (male/female) | 181/281 |
| Race (*n*) | |
| Black/African American | 64 |
| White/Caucasian | 351 |
| Asian/Native Hawaiian/Other Pacific Islander | 35 |

| | |
|---|---|
| More Than One Race | 6 |
| Unknown or Not Reported | 6 |
| Ethnicity (*n*) | |
| Hispanic | 37 |
| Non-Hispanic | 418 |
| Unknown or not reported | 7 |
| Education (years) | 15.34 (*SD*= 1.59) |

Demographic characterization of the HCP Cohort.

**Acute Drug Administration Cohort.** Clinical Trial Name & Number: Brain Networks and Addiction Susceptibility, NCT01924468

Exclusion criteria included, but were not limited to, a history of psychiatric disorders, HIV-positive status, or neurological illnesses. Medications that led to exclusion included, but were not limited to, benzodiazepines, barbiturates, anticonvulsants, antipsychotics, antidepressants, cold medications, and certain herbal supplements (e.g., Kava, Gingko biloba). All procedures were completed at the National Institute on Drug Abuse and were approved by the institutional review board of the National Institutes of Health. Accrual began in August of 2013 and the study completed in September of 2018.

Sample size was determined by power analysis with the key power analysis pertaining to genotypic and fMRI data – target accrual assured that analysis fall into range to detect expected signal changes when accounting for intra-participant variance. This equaled 80 participants total – with 86 actual enrollees to account for drop out. Notably, this sample size was based in part by the genotypic analysis which was not of interest to the present study. The present sample size (N=58) is within standard range for the present study question using fMRI data.

This work was part of a larger trial where the primary goal was evaluating how genotype and catecholaminergic function relates to brain network activity during task and rest. This was assessed by measuring genotype information and measuring how modulation of the dopaminergic and nicotinic system via pharmacological manipulation alters brain network activity at rest and during cognitive tasks. The field has since largely moved past the genotypic analysis which informed the original primary objective, while new fMRI techniques have emerged for understanding brain activity, including retrospective extraction of the sLFO signal. Here, we focused on the effect of pharmacology on the sLFO signal, and assessed how such effects contribute to cognitive task performance.

**Table S4.**

**Demographic Characteristics of Acute Drug Administration Cohort.**

| Demographics | |
|---|---|
| | **Healthy Controls (N = 58)** |
| Age (years) | 31.17 (*SD* = 9.37) |
| Sex (male/female) | 17/41 |
| Race (*n*) | |
| Black/African American | 10 |
| White/Caucasian | 37 |
| Asian | 5 |
| More Than One Race | 6 |
| Ethnicity (*n*) | |
| Hispanic | 9 |
| Non-Hispanic | 48 |
| Unknown or not reported | 1 |
| Education (*n*) | |
| Less than High School | 2 |
| High School Complete/GED | 8 |
| Some / Partial Post-High School | 21 |
| College Graduate/Bachelor's Degree | 18 |
| Master's Degree | 5 |
| Professional Degree (MD, JD, Ph.D.) | 4 |

Demographic characterization of the Acute Drug Administration Cohort.

**fMRI data acquisition**

**Cue Reactivity Cohort.** All participants smoked one of their own cigarettes 1.5 hours prior to the start of MRI scanning to standardize the time since a cigarette was last smoked. Data were acquiring on a Siemens Prisma 3T Scanner (Erlangen, Germany) with a 64-channel head coil. The total scanning session included a 6-minute resting-state acquisition prior to the five, 5-minute runs of the cue reactivity task. Functional scans acquisition used the following parameters: TR = 0.72 s, TE = 30 ms, slices = 66, phase encode direction posterior to anterior, flip angle = 66$^o$, voxel size = 2.5 mm isotropic, GRAPPA acceleration factor = 2, multi-band acceleration factor = 6. Following the functional scans, structural images were acquired with the following parameters: TR = 2.53 s, TE1 = 3.3 ms, TE2 = 6.98 ms, TE3 = 8.79 ms, TE4 = 10.65, flip angle = 7$^o$, resolution= 1.33 x 1.0 x 1.0 mm.

Standard preprocessing was conducted using tools from the FMRIB software library (FSL) (www.fmrib.ox.ac.uk/fsl), including motion correction with MCFLIRT, brain extraction using BET, slice timing correction, spatial smoothing with a full-width half-maximum Gaussian kernel = 6 mm and high-pass filtering at 0.01 Hz.

**Chronic Nicotine Use and Matched Controls Cohort.** On the sated visit day, the participants in the nicotine-smoking group were instructed to smoke *ad lib* prior to MRI scanning. During the abstinence scan day, participants had abstained from smoking for at least 48 hours and as confirmed with a significant reduction in carbon monoxide levels (2.63 $\pm$ 1.90 ppm; $t(64) = 14.79$, $p < 0.001$). Data were acquired on a Siemens Trio 3T Scanner (Erlangen, Germany) using a 12-channel head coil. The total scanning session included an 8-minute resting state MRI scan. Functional scan acquisition used the following parameters: TR = 2.0 s, TE = 27 ms, flip angle = 78°, resolution = 3.44 mm x 3.44 mm x 4 mm. Following the resting state scan, structural images were acquired using the following parameters: TR = 1.9 s, TE= 3.51 ms, flip angle = 9°, resolution: 1.0 x 1.0 x 1.0 mm.

To preprocess the data, raw MRI DICOM data were converted to the standard BIDS structure with bidskit (https://github.com/jmtyszka/bidskit). Data were preprocessed using fMRIPrep (v.23.1.4, https://fmriprep.org/en/23.1.4/), which included discarding the first 2 volumes to account for the T1 non-equilibrium, slice timing correction, volume registration, head motion estimation, and spatial normalization to MNI space.

**HCP Cohort.** The opensource HCP minimal preprocessed resting-state fMRI data was used in this work. Data were acquired on a Siemens 3T Skyra scanner (Erlangen, Germany) modified to achieve a maximum gradient strength of 100 mT $m^{-1}$ using a 32-channel head coil. Participants underwent two resting-state scans occurring on different days. Data were acquired across four, 14.4-minute resting state runs—two runs were completed during one visit and two in a separate visit. Within each session, oblique axial acquisitions alternated between phase encoding in a right-to-left (RL) direction during one run and phase encoding in a left-to-right (LR) direction during the other run. Functional scan acquisition used the following parameters: TR = 0.72 s, TE = 33.1 ms, flip angle = 52°, field of view = 280 x 180 $mm^2$, matrix = 140 x 90, echo spacing = 0.58 ms, bandwidth = 2,290 Hz $px^{-1}$. Slice thickness was set to 2.0 mm, 72 slices and 2.0 mm isotropic voxels, multiband acceleration factor = 8.

**Acute Drug Administration Cohort.** All participants underwent scanning across three drug conditions, which were administered on separate days in a double-blind placebo-controlled design: placebo/placebo, placebo/nicotine, and placebo/methylphenidate. The drug doses were 7 mg of transdermal nicotine (patch) and 20 mg oral methylphenidate. Each scanning session was identical and took place at the peak drug effect: 2 hours post-nicotine and 1 hour post-methylphenidate/placebo, timed according to absorption rates to ensure high and stable plasma levels of medication during the scan. To maintain blinding, participants received either drug or placebo twice on a given scan day—once at the nicotine timepoint and once at the methylphenidate time point. On the placebo scan day, both timepoints involved the administration of placebo. Participants only received one active drug on each study day and placebo at the other timepoint. If participants were discontinued, slots were reused to maintain the balanced randomization. The study physician generated this randomization and assigned participants to interventions. The study physician and lead associate investigators were responsible for enrolling participants. All 58 subjects included in our analysis completed all sessions and did not having missing data.

Data were acquired on a Siemens Trio 3T scanner (Erlangen, Germany) using a 12 channel head coil. The total scanning session included an 8-minute resting-state acquisition prior to the three, approximately 11-minute blocks of the Multi-Source Interference Task (MSIT). Functional scan acquisition used the following parameters: TR = 2 s, TE = 27 ms, flip angle = 78$^o$, resolution = 3.4375 mm x 3.4375 mm x 4 mm. Following the resting state scan, structural images using were acquired using the following parameters: TR = 1.9 s, TE= 3.51 ms, flip angle = 9$^o$, resolution: 1.0 x 1.0 x 1.0 mm.

To preprocess the data, raw MRI DICOM data were converted to the standard BIDS structure with bidskit (https://github.com/jmtyszka/bidskit). Data were preprocessed using fMRIPrep (v.22.1.0, https://fmriprep.org/en/22.1.0/), which included discarding the first 2 volumes to account for the T1 non-equilibrium, slice timing correction, volume registration, head motion estimation, and spatial normalization to MNI space.

### Supplemental Figures

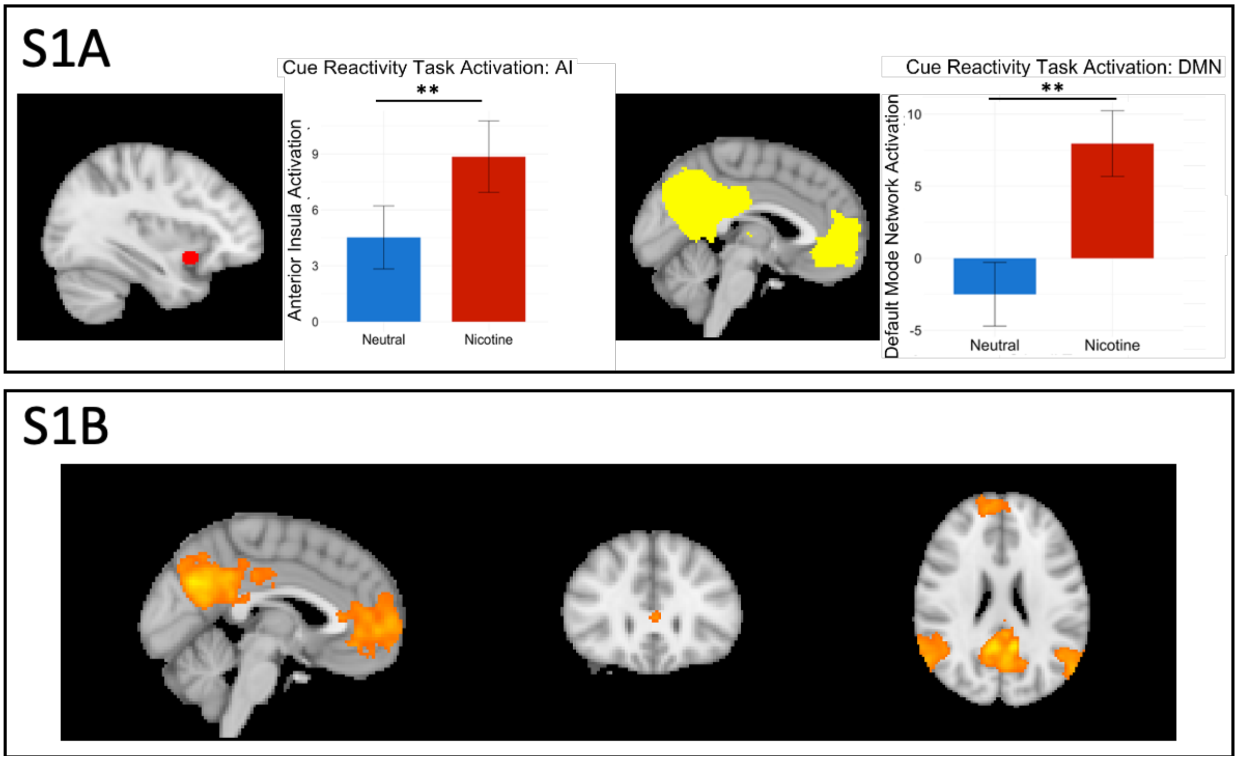

**Supplemental Figure 1: Cue reactivity in the insula and Default Mode Network using BOLD data with the sLFO removed.** S1A: Depict the regions of interest (Adapted from Murray *et al.*, 2025 ) defined for the anterior insula and default mode network, which overlap with known whole brain activation patterns (S1B). There was no relationship between the brain-wide sLFO across the cue reactivity task and the smoking > neutral activation within the DMN (r = -0.18; 95% CI [-0.41, 0.07]; $p = 0.259$) or insula (r = -0.10; 95% CI [-0.34, 0.15]; $p = 0.417$). *** $p < 0.01$

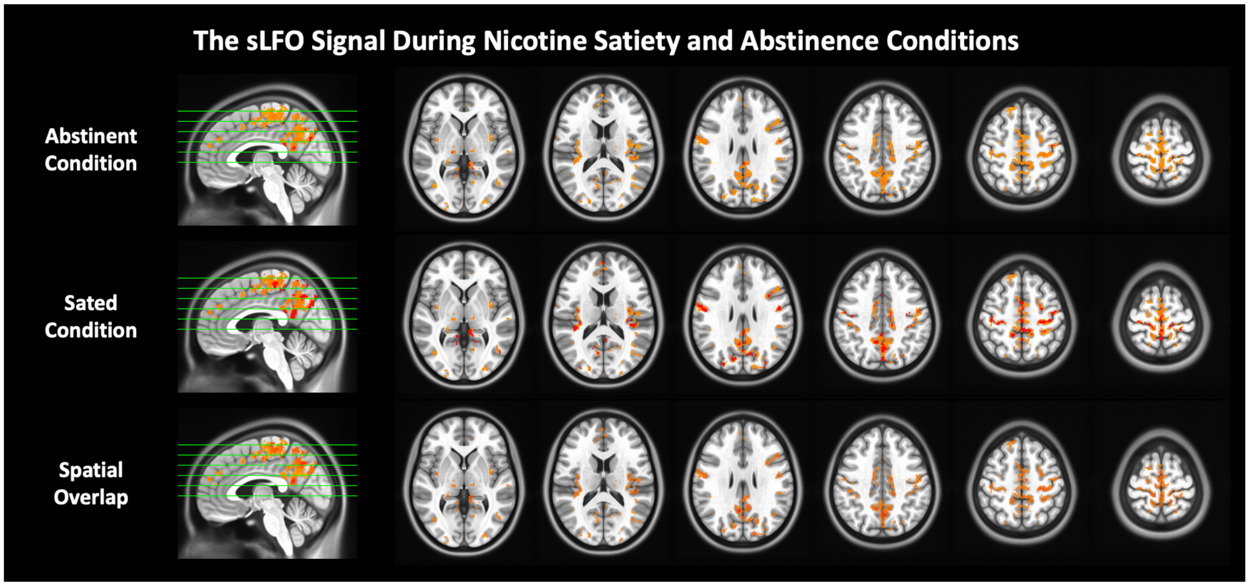

**Supplemental Figure 2: Brain regions showing significant increases in sLFO signal during abstinence relative to satiety in those who use nicotine chronically.** The sLFO signal significantly increased during abstinence (top row) relative to satiety (middle row) in regions between the superior sagittal sinus, inferior sagittal sinus, medial primary motor cortex (mM1), and precuneus ($p < 0.001$, cluster size > 160 mm³; bottom row).

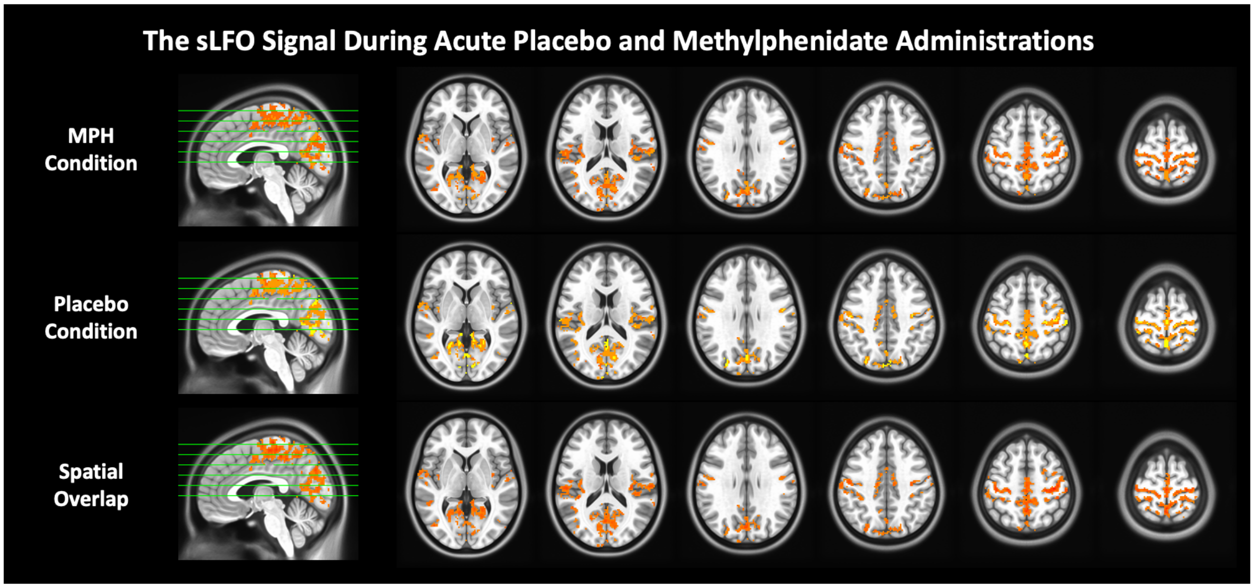

**Supplemental Figure 3: Brain regions showing significant decreases in sLFO signal during acute methylphenidate administration relative to placebo at rest.** The sLFO signal significantly decreased under methylphenidate (MPH; top row) compared to placebo (middle row) in regions between the superior sagittal sinus, inferior sagittal sinus, medial primary motor cortex (mM1), precuneus, and cuneus ($p < 0.001$, cluster size > 160 mm³; bottom row).
