## Supplementary material for "Systemic physiological “noise” in fMRI has clinical relevance": Human Connectome Project (HCP) Cohort Accession Numbers

| Accession Number |
| --- |
| 100206 |
| 100307 |
| 100610 |
| 101309 |
| 102311 |
| 102715 |
| 102816 |
| 103414 |
| 103818 |
| 104012 |
| 105014 |
| 105923 |
| 106016 |
| 106521 |
| 106824 |
| 107422 |
| 108222 |
| 108323 |
| 108525 |
| 109325 |
| 109830 |
| 110411 |
| 111009 |
| 111413 |
| 111716 |
| 112314 |
| 112920 |
| 113215 |
| 113619 |
| 114217 |
| 114621 |
| 114823 |
| 115017 |
| 115320 |
| 115825 |
| 117021 |
| 117122 |
| 117324 |

118528
118831
119126
120111
120515
121416
122317
122822
123117
123420
123521
123723
123925
124422
124826
125424
126426
126628
127327
127630
127731
127832
128127
128632
128935
129028
129129
129331
130013
130518
130619
130821
130922
131217
132017
132118
133019
133625
134223

135225
135528
135932
136833
137128
137431
137532
137633
138231
138534
139435
139637
141422
142828
143224
144125
144731
144832
145127
145834
146129
146937
147737
148133
149337
149741
150625
151223
151425
151728
151829
152427
153025
153126
153227
153732
154330
154431
154532

154734
154835
155635
155938
156435
156536
156637
157336
157437
158035
159441
161630
162026
162228
162733
163129
163432
164030
164131
164939
165638
166640
167238
168139
168341
169444
169545
170631
171532
172029
172332
172433
172534
172938
173334
173435
173536
173738
173839

173940
174437
174841
175136
175439
175540
176441
176542
176845
177746
178142
178243
178849
178950
179245
179346
180230
180735
182032
182739
185139
185846
185947
186040
187143
188448
188549
189349
191235
191942
192035
192540
192843
193239
193845
194140
194443
195041
196144

196346
197348
198047
198350
198653
199352
200311
200614
201414
201515
201818
202113
203418
204420
204622
205119
205725
205826
206323
206525
206828
208630
209329
209935
210415
211114
211619
211821
212015
212318
213421
213522
214221
214423
224022
228434
231928
233326
237334

245333
246133
248339
250427
250932
263436
281135
283543
285446
286347
286650
287248
289555
293748
297655
298051
304020
304727
305830
307127
308129
309636
316633
318637
320826
321323
322224
325129
329844
330324
334635
346137
349244
350330
356948
360030
365343
366446
368551

368753
376247
378756
378857
381543
382242
385046
390645
391748
392750
393247
393550
394956
395251
395756
397760
397861
406432
412528
415837
419239
422632
432332
436845
441939
448347
453441
456346
459453
463040
469961
475855
495255
499566
510326
512835
513130
513736
516742

518746
522434
529549
529953
530635
531940
540436
547046
552544
553344
555348
555651
558657
562446
566454
567052
567961
568963
570243
571144
572045
573249
573451
576255
580650
581450
585862
586460
588565
594156
598568
601127
604537
614439
615441
615744
616645
620434
622236

626648
628248
634748
638049
645551
647858
654350
656657
657659
660951
662551
668361
671855
673455
675661
680250
680452
683256
686969
693764
702133
709551
713239
715647
715950
724446
728454
731140
732243
734045
744553
748662
749058
749361
753251
756055
757764
759869
761957

765864
770352
773257
783462
784565
788674
788876
789373
792564
793465
803240
815247
816653
818455
826353
826454
828862
833148
833249
835657
841349
849264
852455
856766
856968
859671
861456
865363
871762
872562
872764
873968
877168
877269
878776
878877
880157
887373
888678

889579
891667
894067
894673
894774
899885
901139
904044
912447
917255
917558
919966
922854
926862
930449
932554
933253
937160
942658
943862
951457
952863
955465
958976
965367
966975
969476
970764
979984
984472
987074
992673
993675
996782
